# Prediction of SARS-CoV-2 Omicron Variant Immunogenicity, Immune Escape and Pathogenicity, through Analysis of Spike Protein-specific Core Unique Peptides

**DOI:** 10.1101/2021.12.26.21268418

**Authors:** Vasileios Pierros, Evangelos Kontopodis, Dimitrios J. Stravopodis, George Th. Tsangaris

**Affiliations:** Proteomics Research Unit, Biomedical Research Foundation of the Academy of Athens; 11527 Athens, Greece; Section of Cell Biology and Biophysics, Department of Biology, School of Science, National and Kapodistrian University of Athens; 15701 Athens, Greece

**Author notes:** Correspondence (GThT). These authors contributed equally to this work (Vasileios Pierros, Evangelos Kontopodis).

**Keywords:** Core Unique Peptide, COVID-19, Immune escape, Infectiveness, Mutation, Omicron variant, Pathogenicity, SARS-CoV-2, Spike protein, Uniquome

## Abstract

The recently discovered Omicron variant of the SARS-CoV-2 corona virus has raised a new, global, awareness, since it is considered as a new variant of concern from all major health organizations, including WHO and ECDC. Omicron variant is characterized by 30 amino acid changes, three small deletions and one small insertion in the Spike protein. In this study, we have identified the Core Unique Peptides (CrUPs) that reside exclusively in the Omicron variant of Spike protein and are absent from the human proteome, thus creating a new dataset of peptides named as C/H-CrUPs. Furthermore, we have analyzed their protein locations and compared them with the respective ones of Alpha and Delta SARS-CoV-2 variants. In Omicron, 115 C/H-CrUPs were generated and 119 C/H-CrUPs were lost, almost four times as many compared to the other two variants. From position 440 to position 508, at the Receptor Binding Motif (RBM), 8 mutations were detected, resulting in the construction of 28 novel C/H-CrUPs. Most importantly, in Omicron variant, new C/H-CrUPs carrying two or three mutant amino acids were produced, as a consequence of the accumulation of multiple mutations in the RBM. Remarkably, these Omicron-derived C/H-CrUPs that bear several mutated amino acids could not be recognized in any other viral Spike variant. We suggest that virus binding to the ACE2 receptor is facilitated by the herein identified C/H-CrUPs in contact point mutations and Spike-cleavage sites, while the immunoregulatory NF9 peptide is not detectably affected. Taken together, our findings indicate that Omicron variant contains intrinsic abilities to escape immune-system attack, while its mutations can mediate strong viral binding to the ACE2 receptor, leading to highly efficient fusion of the virus to the target cell. However, the intact NF9 peptide suggests that Omicron exhibits reduced pathogenicity compared to Delta variant.

## Introduction

SARS-CoV-2 virus has been presented with an ability of high mutagenesis frequency, hitherto producing 63 different variants with 39 of them being considered as the most predominant forms, including Delta, the dominant variant of the 4^th^ pandemic wave (1). Recently, a new variant, the Omicron (B.1.1.529), was identified in South Africa. Omicron is characterized by 30 amino acid changes, three small deletions and one small insertion in Spike protein, as compared to the original virus, with 15 of them residing in the Receptor Binding Domain (RBD) from 319 to 541 amino acid residues (2).

To elucidate the SARS-CoV-2 virus-host organism interactions, we have designed a novel bioinformatics approach to analyze the Core Unique Peptides (CrUPs) of the SARS-CoV-2 virus against the human proteome (C/H-CrUPs) (1). C/H-CrUPs represent a completely new set of peptides, which are the sortest in length peptides in viral proteome that do not exist in human proteome. Based on their properties, the viral C/H-CrUPs could advance our knowledge regarding virus-host interactions, immune-system response(s), infectiveness and pathogenicity of the virus, while, most importantly, they can be used as antigenic and diagnostic peptides, and likely druggable targets for successful therapeutic treatments.

In the present study, we have identified, cataloged and analyzed the Omicron-specific C/H-CrUPs, in order to illuminate the mechanisms controlling infectivity, immune escape and pathogenicity of the new variant.

## Materials and Methods Methods

For our recent studies (1, 3), we have developed a bioinformatics tool that can extract from a given proteome the Core Unique Peptides (CrUPs) (thus creating its Uniquome) (Supp. Fig. 1). We have expanded this tool by introducing a new feature that can extract the CrUPs of each individual protein of a given proteome (target) versus the proteins of a reference proteome. This new feature, like the initial implementation, will split each protein in the target proteome to all possible peptides of length minimum (4 amino acids) to length maximum (100 amino acids) and search them against the reference proteome. Each search will exclude all peptides that contain a smaller peptide already identified as CrUP (Supp. Fig. 2).

For the present study, we have engaged this new feature of our tool. We have created a “custom” proteome consisting of sequences from all variants of the SARS-CoV-2 Spike proteins and used it as the target versus the human proteome. The tool produces as output the C/H-CrUPs per protein of the target proteome, thus revealing the CrUPs for each Spike variant versus the human proteome.

Once we have the desired data, we run a meta-analysis to identify how many C/H-CrUPs remain the same, or are added or lost on each variant versus the wild-type Spike protein. For this analysis, initially we take the identified C/H-CrUPs of the wild-type sequence and check their presence against the respective C/H-CrUPs of the other variants. We only care for the amino acid sequence and not the position this could be found within the protein. If the sequence is found, then we consider the peptide to be the same, otherwise we consider it to be lost on the examined variant.

Next, we analyze the identified C/H-CrUPs of each variant versus the wild-type sequence. If the peptide is detected only on variant’s C/H-CrUPs, then we consider it as added. This meta-analysis also provides us with the position of each C/H-CrUP within the Spike protein, which we have used to determine the area (i.e. RBD, or S-cleavage site) they reside in.

## Databases

All proteomes and proteins were obtained from Uniprot. SARS-CoV-2 wild-type and variant sequences, and mutations were obtained from Stanford COVID Database (https://covdb.stanford.edu/page/mutation-viewer/).

## Results and Discussion

### Mapping the C/H-CrUPs landscape of Spike protein of SARS-CoV-2 Omicron variant

SARS-CoV-2 virus seems to be highly mutated, producing so far more that 60 distinct variants. Hitherto, the highest pathogenic form is the Delta variant (B.1.617.2), with 10 different sub-variants. Recently, a novel variant, called Omicron, has been identified. It is characterized by 30 amino acid changes, three small deletions and one small insertion in the Spike protein area, as compared to the wild-type viral respective sequence (Supp. Fig. 3) (2). Out of these genetic changes, 15 reside in the Receptor Binding Domain (RBD) from amino acid position 318 to 541 and two are located around the S-cleavage site(s) (Supp. Fig. 3).

Advanced bioinformatics analysis of the Omicron variant Spike protein showed that it contains 983 C/H-CrUPs, a number that is comparable to the one of wild-type Spike protein (987 C/H-CrUPs) and to the mean ± SD value of Spike protein-specific C/H-CrUPs (983±2 C/H-CrUPs) (Table 1). Omicron variant Spike protein contains 34 mutations in total, which is the highest number of identified mutations among all virus variants.

**TABLE 1.**
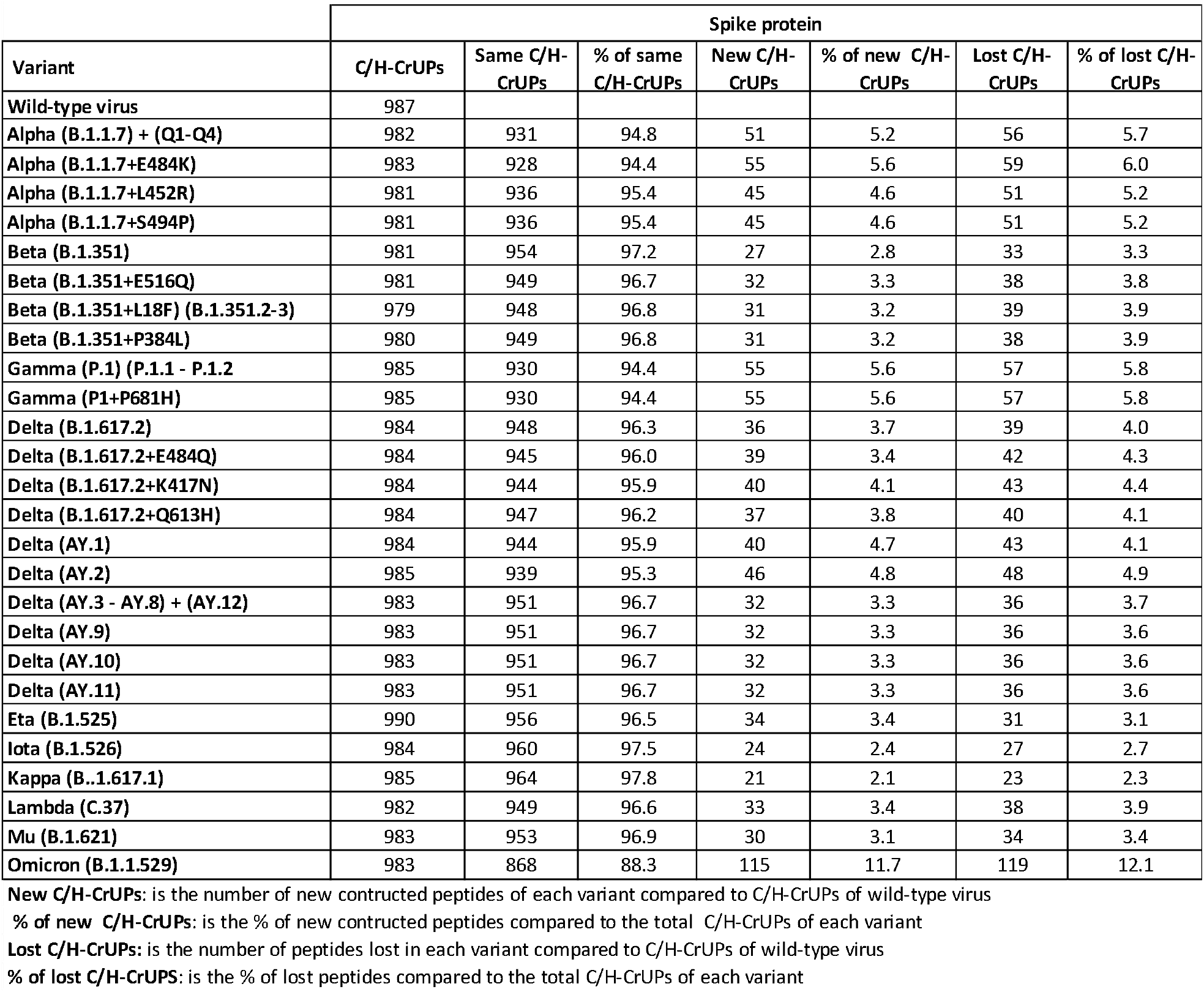
SARS-CoV-2 Spike protein C/H-CrUPs across variants, as compared to the wild-type virus respective sequence

These mutations seem to have a dramatic effect on Spike protein C/H-CrUPs map. Compared to the wild-type Spike sequence, we found that 115 (new) C/H-CrUPs were created and 119 C/H-CrUPs were lost, almost twice as many when compared to the Alpha variant (51 and 56 C/H-CrUPs, respectively), and almost four times as many, compared to other variants (Table 1). The distribution of these new C/H-CrUPs shows that their majority carry 6 amino acids in length (Supp. Fig. 4).

### Omicron-specific C/H-CrUPs that belong to the Receptor Binding Domain

SARS-CoV-2 belongs to the β coronavirus group, which uses the plasma membrane receptor of Angiotensin-Converting Enzyme 2 (ACE2) to recognize and bind to the target cell (Walls *et al*., 2020; Wang *et al*., 2020). The viral Spike protein attaches to ACE2 receptor by a Receptor Binding Domain (RBD) defined from amino acid position F318 to F541 (4). The amino acid residues from W436 to Q506 inside RBD shape the Receptor Binding Motif (RBM), which carries 11 contact positions with ACE2 (5). The RBD region has received great attention, as it seems to be a major target of antibodies against the virus and other therapeutic interventions (6-8).

In the RBD region, Omicron variant carries 15 mutations, 10 of which are identified in the RBM area (Fig. 1A). This results in identification of the highest number of newly constructed C/H-CrUPs in the RBD/RBM region, as compared to all other previous virus variants examined (Supp. Table 1). Table 2, describes all the new, herein identified, C/H-CrUPs of Omicron variant in Spike’s RBD region, in comparison to Alpha and Delta variants, which represent two of the most predominant variants of the virus in human populations. Hence, it proved that, in contrast to Alpha and Delta variants, at the end of Omicron variant RBM area, from 440 to 508 amino acid position, 8 novel mutations were identified, resulting in production of 28 new C/H-CrUPs. The most important finding is that in Omicron variant, for the first time, new C/H-CrUPs including two or three mutant amino acids were generated, with the peptides “*QAGN*K*P*”, “*N*K*PCN*”, “*LK*SYS*F*” and “*K*SYS*FR*\*” being characteristic examples, as a result of the accumulation of multiple mutations in the positions 440, 446, 477, 478 and 493-505. These novel C/H-CrUPs that contain several mutated amino acids could not be found in any other virus variants till today. Taking into consideration recent data about virus infectivity, the multimutated, new, C/H-CrUP collection seems to change radically the structure and the epitope regions of end positions of the RBM area, in Omicron variant, causing serious compromise of its antigenic capacity and facilitating the immune escape of the virus (9).

**Figure 1.**
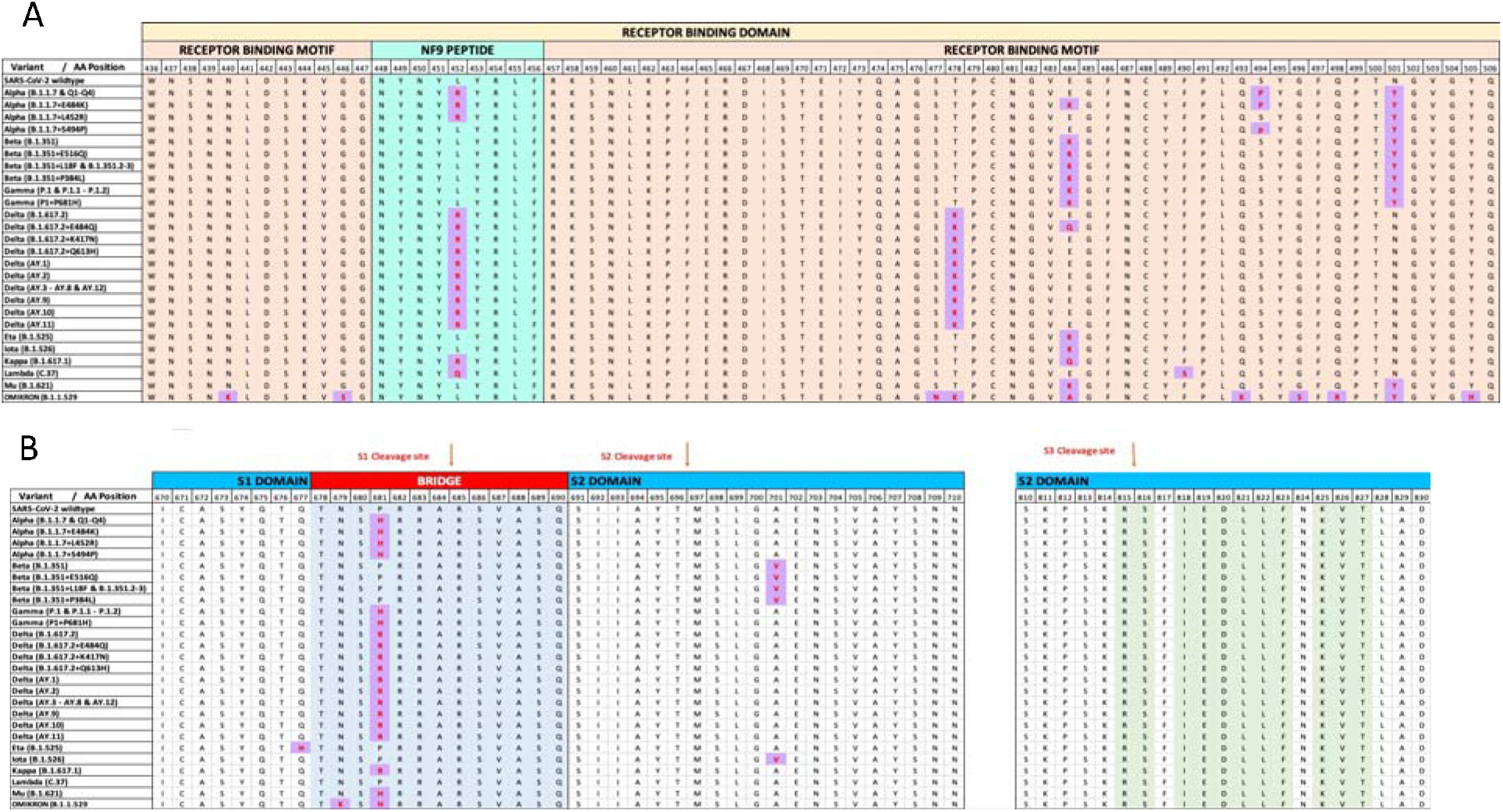
Presentation of the SARS-CoV-2 mutations in different virus variants. **A**) The mutations of the Receptor Binding Motif (RBM) which included in the Receptor Binding Domain (RBD) were presented. **B)** The mutations around the Spike cleavage sites were presented. Purple blocks marked the point mutations sites in the variants, green color indicate the Universal Peptides of the spike proteins from Fig. S2. Yellow color mark the Receptor-Binding Domain of spike protein to ACE2, pink color mark the Receptor-Binding Motif, cyan mark the NF9 peptide and light blue mark the Bridge between S1 and S2 domain. Red arrows indicate the cleavage sites. With different colors in the upper side of the alignment, the different domains of the spike protein are marked.

**Table 2.**
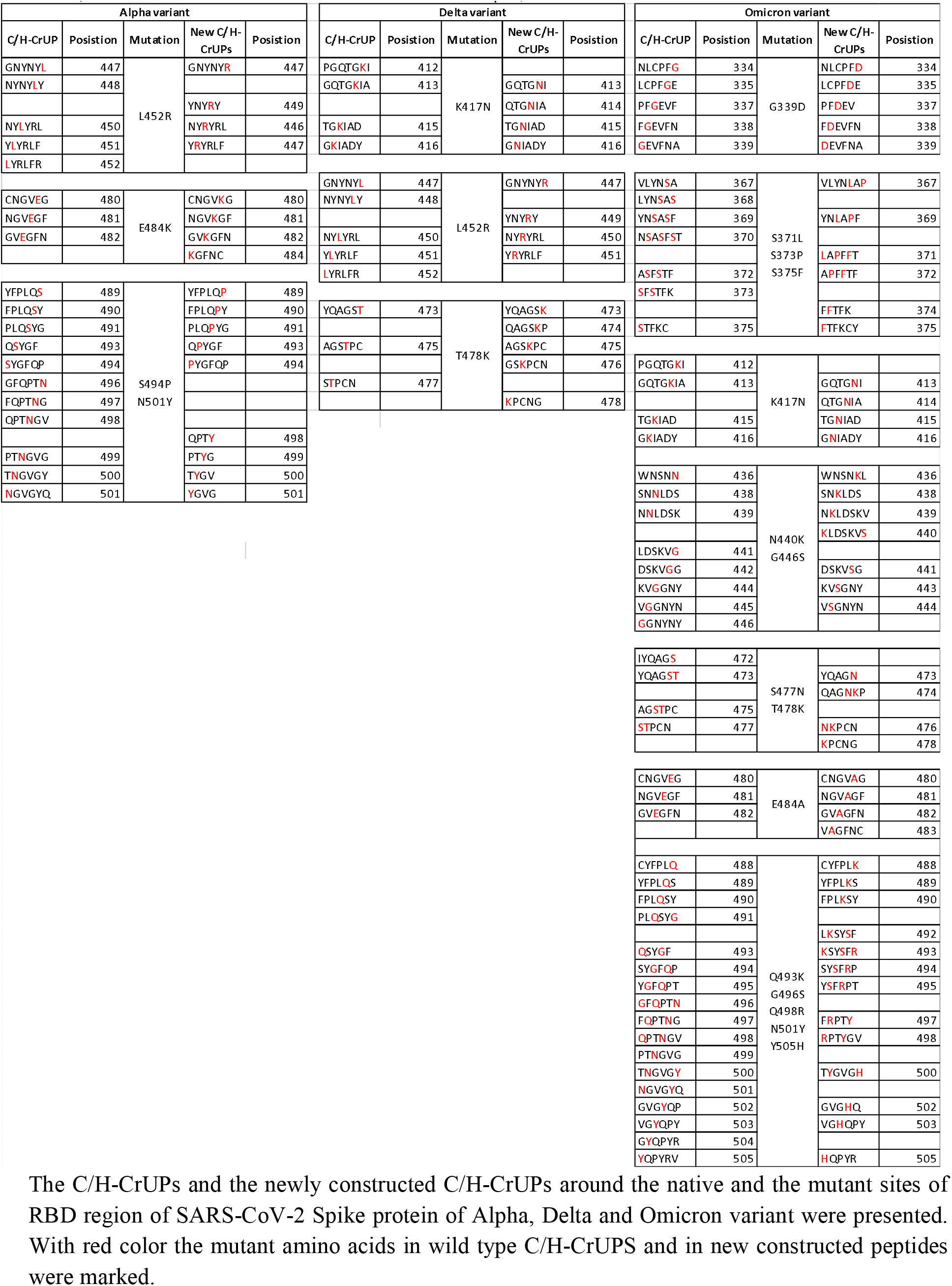
C/H-CrUPs constructed aroud the mutations in RBD of Alpha, Delta and Onicron SARS-CoV 2 variants

Remarkably, RBM area contains 11 out of the 12 contact points of viral Spike protein with the ACE2 cellular receptor. Among them, 7 contact points remained intact, while 4 mutations in positions Q493K, Q498R, N501Y and Y505H were identified, resulting in the construction of 17 new C/H-CrUPs (Table 3). N501Y mutation was found to be a major determinant of increased viral transmission, due to improved binding affinity of Spike protein to ACE2 cellular receptor (12). These findings indicate that virus binding to ACE2 receptor is notably affected by C/H-CrUP-specific mutations that can likely strengthen the Spike-ACE2 protein-protein interaction(s). Interestingly, an important amino acid sequence in RBM area is the “*NYNYLYRLF*” peptide (from 448 to 456 position). This Tyrosine (Y)-enriched peptide carries two contact sites (Y449 and Y453) and it is known as the NF9 peptide (10). It seems to affect antigen recognition, by being an immunodominant HLA*24:02-restricted epitope identified by CD8^+^ T cells. Of note, NF9 presents immune stimulation activity, and increases cytokine production derived from CD8^+^ T cells, such as IFN-γ, TNF-α and IL-2 (11). In contrast to Delta, in Omicron variant the NF9 amino acid content is not changed by any mutation detected, thus suggesting that the NF9 peptide could induce an early immune system activation and efficient cytokine production, leading to a faster immune response, and thus reducing SARS-CoV-2 virus pathogenicity.

**Table 3.**
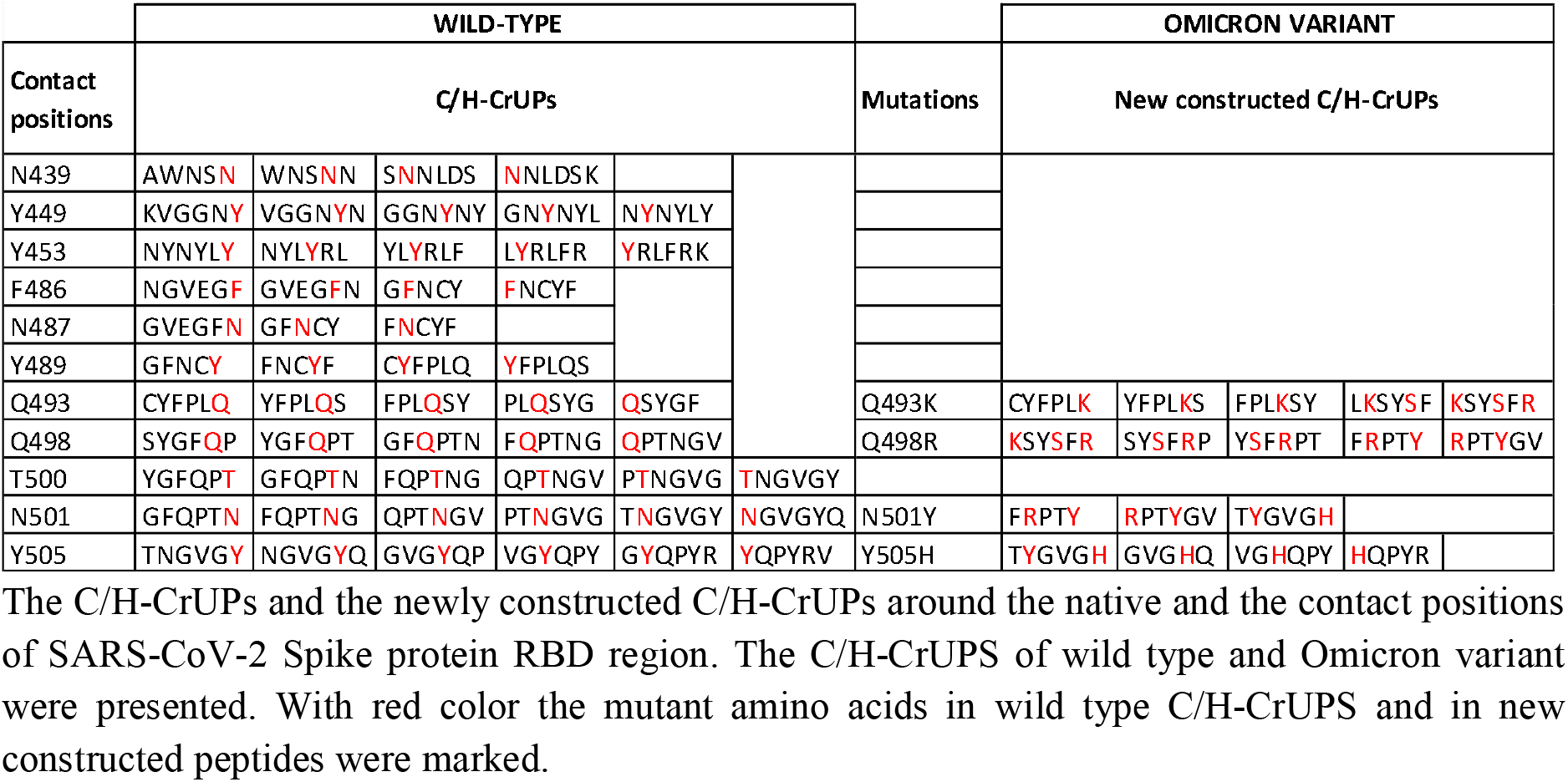
C/H-CrUPs around SARS-CoV-2 RBD contatcs positions

### C/H-CrUPs altered architecture around the Spike-cleavage site(s) of Omicron variant

The molecular mechanism of Spike protein’s proteolytic activation has been shown to play a crucial role in the selection of host species, virus-cell fusion, and viral infection of human lung cells (13-15). Spike protein [SPIKE_SARS2 (P0DTC2)] contains three cleavage sites (known as S-cleavage sites) crucial for the virus fusion to the host cell: the R685↓S and R815↓S positions that serve as direct targets of the Furin protease, and the T696↓M position that can be recognized by the TMPRSS2 protease (16-18).

In these cleavage sites Omicron variant carries only the critical mutation P681H, which also appears in the Alpha variant (Fig. 1B). Strikingly, in contrast to Delta variant, which contains the P681R mutation, the P681H mutation constructs several new C/H-CrUPs in the Alpha and Omicron variants, thus indicating their dispensable contribution to virus fusion to the host cell (Table 4).

**TABLE 4.** C/H-CrUPs arround the Spike protein cleavage sites.

| Cleavage Site | Mutation | Variant | Position | New C/H-CrUPs |
| --- | --- | --- | --- | --- |
| R<sup>685</sup>↓S | P681R | Delta | 680 | SRRRAR↓S |
|  | P681H | Alpha Omicron | 677 | QTNSH |
|  |  |  | 678 | TNSHR |
|  |  |  | 680 | SHRRAR |
| T<sup>696</sup>↓M | A701V | Beta | None |  |
| R<sup>815</sup>↓S | None | None | None |  |

The C/H-CrUPs around the mutant positions of Spike protein cleavages sites were presented. ↓ indicates the protein cleavage position.

## Conclusions

Analysis of C/H-CrUP landscapes in heavily mutated SARS-CoV-2 Omicron variant Spike protein unveiled that the Omicron variant, by the generation of novel multimutated C/H-CrUPs could escape the immune system defense mechanisms, while these C/H-CrUP-specific mutations could facilitate the more efficient virus binding to the ACE2 cellular receptor, and to the more productive fusion of the virus to the host cell. Most importantly, in contrast to Delta variant, in Omicron variant the intact NF9 peptide, which has a known immunostimulatory effect, suggests that Omicron variant exhibits reduced pathogenicity, as compared to the Delta one.

## Data Availability

All data produced in the present work are contained in the manuscript

## Author contributions

Conceptualization: VP, EK, GThT; Methodology: VP, EK; Investigation: VP, EK, GThT; Visualization: EK, DJS, GThT; Supervision: GThT; Writing - original draft: DJS, GThT; Writing - review & editing: DJS, GThT.

## Data and materials availability

All data are available in the main text or in the supplementary materials.

