## SUPPLEMENTARY DATA for "Prediction of SARS-CoV-2 Omicron Variant Immunogenicity, Immune Escape and Pathogenicity, through Analysis of Spike Protein-specific Core Unique Peptides"

**Affiliations:**

SUPPLEMENTARY DATA

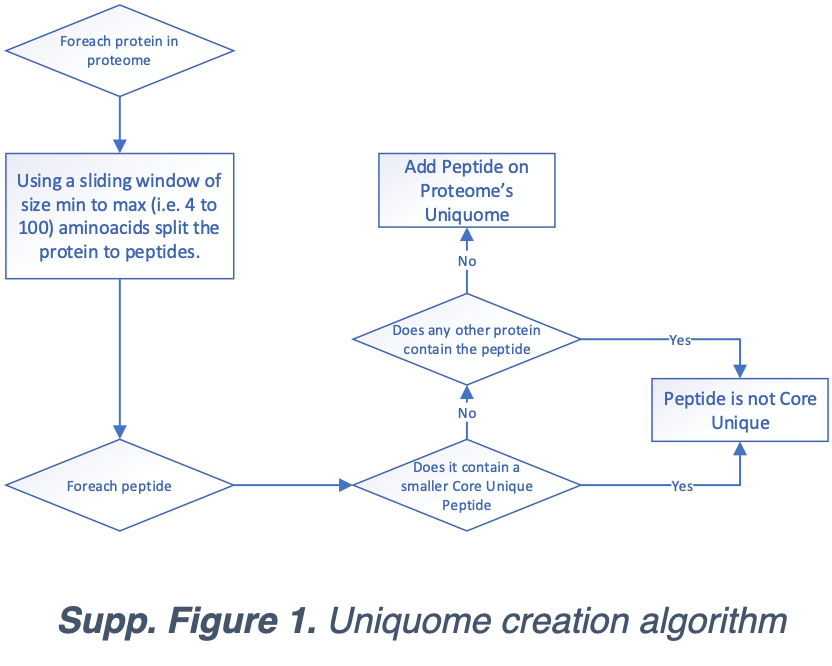

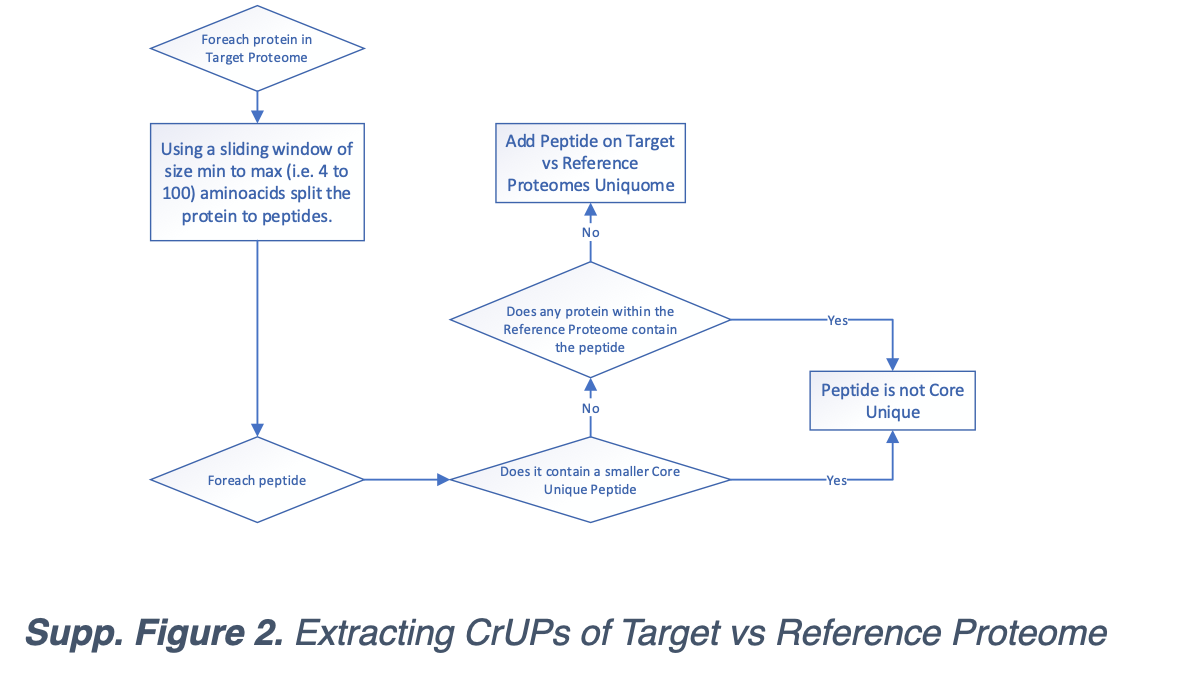

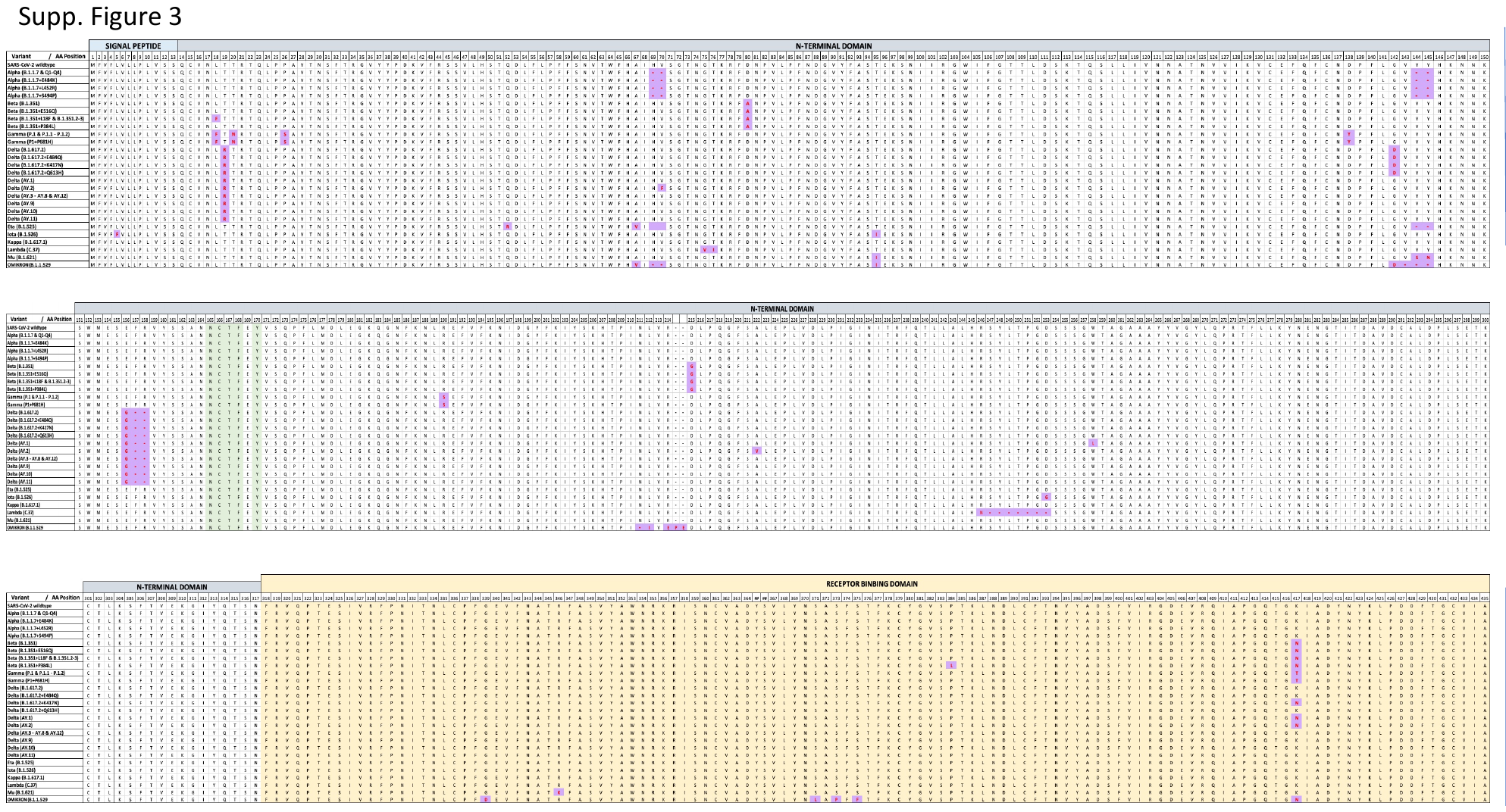

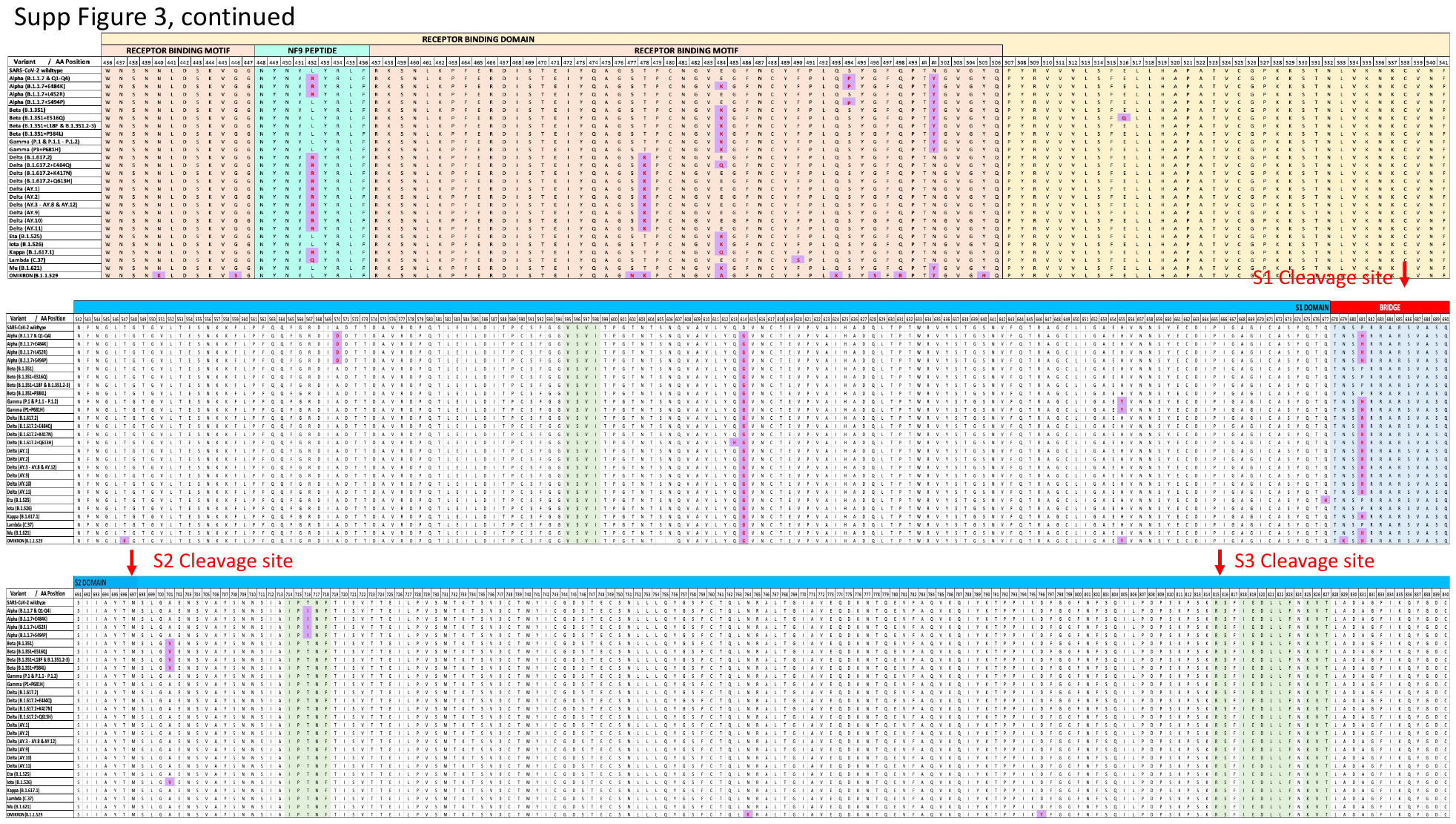

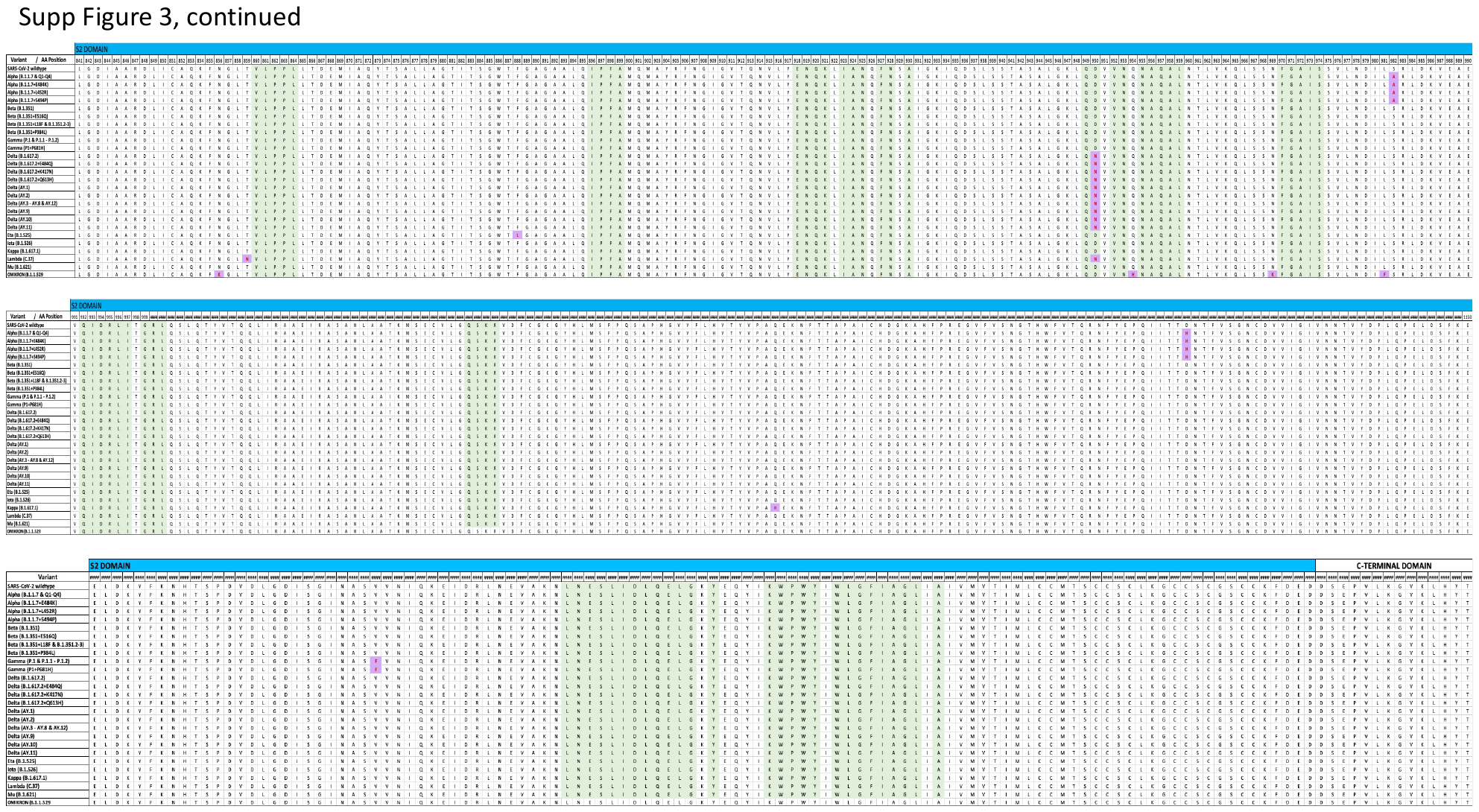

***Supp. Figure 3. Alignment of the SARS-CoV-2 spike protein (SPIKE_SARS2, P0DTC2) of the 26 variants, together* with the wild-type Spike Protein (SPIKE_SARS2, P0DTC2)**. The complete spike sequence alignment. Purple blocks marked the point mutations sites in the variants, green color indicate the Universal Peptides of the spike proteins from Fig. S2. Yellow color mark the Receptor-Binding Domain of spike protein to ACE2, pink color mark the Receptor-Binding Motif, cyan mark the NF9 peptide and light blue mark the Bridge between S1 and S2 domain. Red arrows indicate the cleavage sites. With different colors in the upper side of the alignment, the different domains of the spike protein are marked.

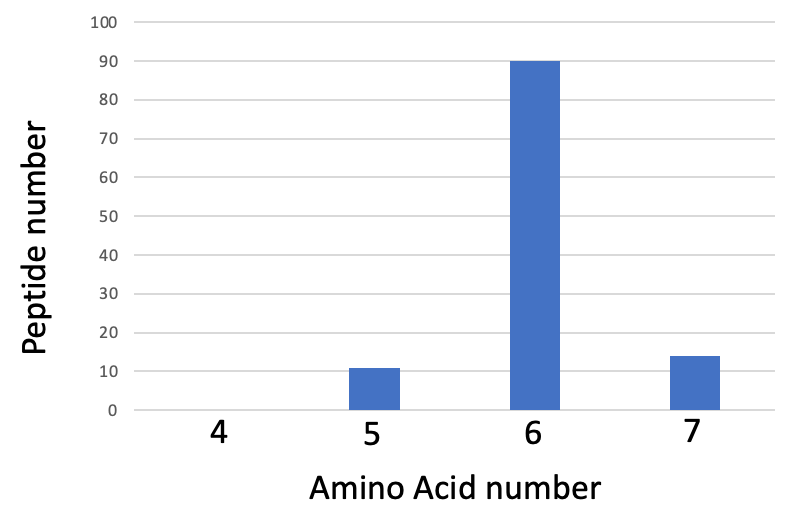

**Supp Figure 4.** Length distribution of Omicron variant Spike protein C/H-CrUPs.

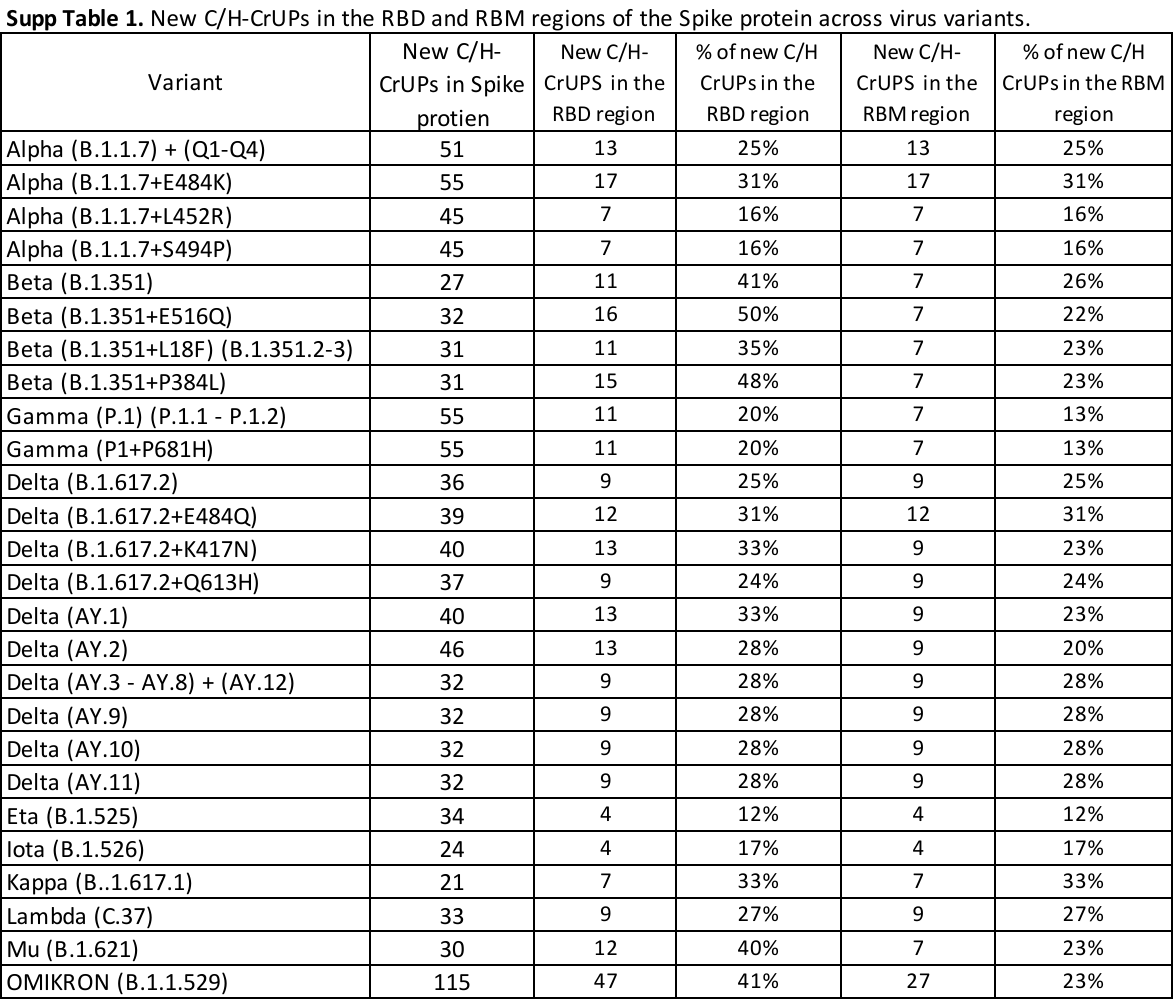
